# Exploring the Performance and Explainability of BERT for Medical Image Protocol Assignment

**DOI:** 10.1101/2023.04.20.23288684

**Authors:** Salmonn Talebi, Elizabeth Tong, Mohammad R. K. Mofrad

**Affiliations:** University of California, Berkeley, Berkeley, CA, USA; Stanford University, Stanford, CA, USA

## Abstract

Although deep learning has become state of the art for numerous tasks, it remains untouched for many specialized domains. High stake environments such as medical settings pose more challenges due to trust and safety issues for deep learning algorithms. In this work, we propose to address these issues by evaluating the performance and explanability of a Bidirectional Encoder Representations from Transformers (BERT) model for the task of medical image protocol assignment. Specifically, we evaluate the performance and explainability on this medical image protocol classification task by fine tuning a pre-trained BERT model and measuring the word importance by attributing the classification output to every word through a gradient based method. We then have a trained radiologist review the resulting word importance scores and assess the validity of the model’s decision-making process in comparison to that of a human. Our results indicate that the BERT model is able to identify relevant words that are highly indicative of the target protocol. Furthermore, through the analysis of important words in misclassifications, we are able to reveal potential systematic errors in the model that may be addressed to improve its safety and suitability for use in a clinical setting.

## 1 Introduction

Machine learning systems are being rapidly adopted for many applications including high-stakes settings such as medical applications [18, 19, 22]. Recent progress with self-attention techniques, and specifically Transformers, have dominated the field of text processing and classification tasks. Large pretrained Transformers have outperformed humans on language understanding tasks such as SuperGLUE [26]. However, many specialized text analysis tasks do not make use of modern machine learning methods. It remains questionable how well existing pretrained models will transfer to large, specialized texts.

In many high-stake applications, such as medicine, law, or security where the main workers are humans trained in specialized tasks, the direct application of these machine learning algorithms, without human oversight, is currently inappropriate. This reason is not only due to accuracy concerns, but also arise from the lack of explanability and trust humans have for the machine learning algorithm. Therefore, in order to implement a machine learning algorithm to help with specialized medical tasks it must not only have human level performance, but also provide trustworthy explanations to the user [10]. Furthermore, model explainability is being driven by laws and regulations which state that decisions from machine learning algorithms must provide information about the logic behind those decisions [1]. In fact, the lack of explainability of ML models often plagues medical artificial intelligence (AI) [8]. For these reasons, in high-stake settings, explainability should be a priority for researchers.

In this study, we focus on the specialized task of identifying medical imaging protocols within text descriptions. Medical imaging plays a crucial role in modern healthcare, allowing physicians to visualize the inside of the human body in order to diagnose and manage various conditions. Clinicians often order radiologic studies, such as magnetic resonance imaging (MRI) or computed tomography (CT), to help answer clinical questions and guide treatment decisions [24].

Typically, when a physician orders an imaging study, he/she will provide a brief description of the indication for the exam outlining the patient’s signs and symptoms, medical history, and any relevant clinical findings. These requests are then sent to the radiologists, who are responsible for reviewing the orders and recommending a radiologic protocol that best addresses the clinical question. A radiologic protocol is a specific set of instructions that defines the type of radiologic exam to be performed on a particular body part, taking into account the patient’s presentation and the expected imaging findings. The protocol may involve different imaging sequences contrast agents, imaging planes, field of view, etc in an MRI exam.

Assigning the appropriate protocol requires a thorough understanding of the radiological appearance of different pathologies, as well as a detailed knowledge of the patients’ clinical presentation and medical history. It also requires familiarity with the types of protocols offered by the institution, as different facilities may have different capabilities and resources. In MRI, accurate protocol assignment is particularly crucial to patient care, as the chosen protocol dictates which sequences are obtained and can impact the quality and diagnostic accuracy of the exam [3, 4].

Traditionally, protocol assignment to each radiologic order is done manually by the radiologists or radiology technologists. This can incur substantial costs to the healthcare system. This tedious task may take up to at least 6% of the radiologists’ time [21]. With increasing radiology orders, an automated process with high throughput and accuracy is desirable to ensure patient care and to avoid radiologists’ burnout. However, given the high stakes of medical tasks, machine learning models must be evaluated for any systematic biases or errors before they can be trusted by clinicians and patients [7]. In order for these models to be used in practice they need to provide valid explanations for how the decisions are made.

To address these problems, we fine-tuned a BERT model using thousands of archived physician orders to learn the medical language used to describe a given radiological exam. Physicians’ orders are generally written poorly, with many typos and grammatical errors. In many cases they are written with a few keywords to try and convey their point. Furthermore, they use terminology that only make sense in the context of human anatomy or physiology. This can pose challenges for existing pre-trained models as there is a distribution shift between the physician’s text and what existing models have been trained on [15]. In addition, we evaluate the model’s ability to provide explanations of its decision based on word importance. A trustworthy algorithm should be able to demonstrate it is making complex decisions using similar rational to a human. For this application, explanation is increasingly complex because the model will need to understand language in the context of human anatomy and physiology.

The main contributions of this study are as follows:

- We fine-tune a pre-trained BERT model using a medical dataset of medical imaging protocol text, and demonstrate that it achieves state-of-the-art performance.
- We employ a gradient-based method called integrated gradients to quantify the contribution that each word in the input text makes to the model’s decision.
- We validate the model’s word importance claims using a technique called erasure.
- We demonstrate that the model is capable of complex decisions in a manner similar to that of a trained radiologist.
- We analyze the model’s mistakes using word importance and identify systematic errors that may pose potential safety risks and need to be addressed before the model can be safely deployed in a clinical setting.

## 2 Data

In order to train a specialized model for medical text classification, we have compiled a new large-scale dataset for image protocol review. This dataset consists of deidentified order entries and assigned protocols for magnetic resonance (MR) neuroradiology studies that were conducted at our institution between June 2018 and July 2021. Each row in the dataset represents a single radiology order and includes the ‘reason for exam’, patient age and gender, and the protocol assigned by the radiologist.

We have excluded orders for spine imaging from this study, as the assigned protocol typically reflects the specific segment of the spine indicated in the order. From the original dataset of 119,093 rows, we removed the most common protocol, ‘routine brain’, as it can be used for a wide range of indications and serves as the default protocol at our institution. The remaining dataset was narrowed down to the 10 most common protocols (Table 1).

**Table 1.**
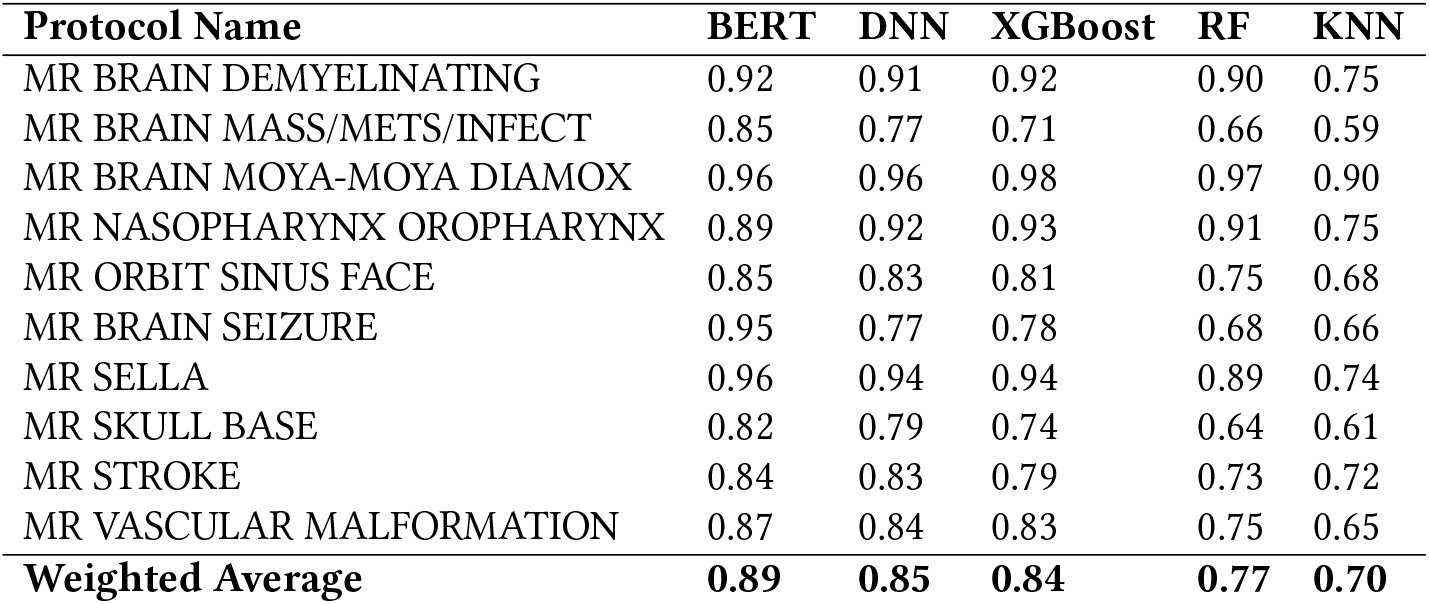
A comparison of imaging protocol F1 scores.

| Protocol Name | BERT | DNN | XGBoost | RF | KNN |
| --- | --- | --- | --- | --- | --- |
| MR BRAIN DEMYELINATING | 0.92 | 0.91 | 0.92 | 0.90 | 0.75 |
| MR BRAIN MASS/METS/INFECT | 0.85 | 0.77 | 0.71 | 0.66 | 0.59 |
| MR BRAIN MOYA-MOYA DIAMOX | 0.96 | 0.96 | 0.98 | 0.97 | 0.90 |
| MR NASOPHARYNX OROPHARYNX | 0.89 | 0.92 | 0.93 | 0.91 | 0.75 |
| MR ORBIT SINUS FACE | 0.85 | 0.83 | 0.81 | 0.75 | 0.68 |
| MR BRAIN SEIZURE | 0.95 | 0.77 | 0.78 | 0.68 | 0.66 |
| MR SELLA | 0.96 | 0.94 | 0.94 | 0.89 | 0.74 |
| MR SKULL BASE | 0.82 | 0.79 | 0.74 | 0.64 | 0.61 |
| MR STROKE | 0.84 | 0.83 | 0.79 | 0.73 | 0.72 |
| MR VASCULAR MALFORMATION | 0.87 | 0.84 | 0.83 | 0.75 | 0.65 |
| **Weighted Average** | **0.89** | **0.85** | **0.84** | **0.77** | **0.70** |

To ensure the accuracy and quality of the data, we performed a thorough review by an experienced radiologist (ET) with 10 years of experience. We also applied standard text preprocessing techniques, such as the removal of redundant fields, handling of missing outputs, and expansion of acronyms, to further clean and organize the data. The final dataset includes 88,000 recorded notes with expert-annotated imaging protocols.

## 3 Methods

This retrospective study was conducted with the approval of the Stanford Institutional Review Board (IRB) and under a waiver of informed consent. The study was approved for collaboration between Stanford University and the University of California, Berkeley.

### 3.1 BERT Fine Tuning

We approach the problem of text classification as predicting the class that corresponds to a given input text. In our dataset, we have 10 possible classes that can be predicted. To achieve this, we fine-tune a pre-trained BERT model using the HuggingFace Transformers library [28].

Before being processed by the BERT encoder, the input data is transformed by passing it through three embedding layers: a token embedding layer, a segment embedding layer, and a position embedding layer. In the token embedding layer, the input sentences are tokenized and each token is transformed into a fixed-dimensional vector representation (e.g., a 768-dimensional vector). Special classification [CLS] and separator [SEP] tokens are also inserted at the beginning and end of the tokenized sentence to serve as input representations and sentence separators for the classification task.

The segment embedding layer is useful for classifying a text when provided with a pair of input texts. The positional embedding layer encodes the relative position of tokens within a sentence using a sinusoidal function. The final input embedding is the sum of these three individual embeddings, which is then passed to the transformer for further processing.

Resemblant to the clinical setting, the number in each protocol is not evenly distributed. More than half of the imaging protocol entries belong to two of the classes. To mitigate this imbalance we up sample the remaining 8 imaging protocols so that the dataset is approximately balanced between all 10 classes of imaging protocols. The data is randomly split into a train, validation and test sets. We have 70% of the protocols make up the train set, 20% make up the validation set, and 10% make up the test set. The validation set was used to perform a hyperparameter grid search. The learning rate was tuned from the range of 1*x* 10^−4^ to 1*x* 10^−6^. During our experiments we found the model would converge after 20 epochs and training for any longer would degrade performance. The model is trained using a single A6000 GPU.

### 3.2 Model Baseline

In order to establish a baseline and compare the performance of our fine-tuned BERT model against traditional machine learning methods, we conducted experiments using several well-known algorithms, namely K-Nearest Neighbors (KNN), Random Forest (RF), XGBoost, and Deep Neural Networks (DNN). These algorithms have been used in previous studies for medical imaging protocol assignment and provide a benchmark to evaluate the effectiveness of our approach.

To implement and evaluate the traditional machine learning methods, we used popular and widely adopted Python libraries for each of the algorithms. For KNN, RF, and XG-Boost, we utilized the scikit-learn library. For the DNN, we employed Keras for building a 1D Convolutional Neural Network (CNN) classifier.

### 3.3 Word Importance

For the purposes of this study, we use the concept of “word importance” as a means of interpreting the model. Word importance quantifies the contribution that each word in the input text makes to the model’s prediction. To calculate word importance, we utilize a gradient-based method called integrated gradients [16, 23].

Integrated gradients exploit the gradient information of the model by integrating first-order derivatives. This method does not require the model to be differentiable or smooth, making it particularly suitable for large and complex models such as Transformers. We use integrated gradients to accurately estimate the importance of individual words within an input sentence.

The integrated gradients method can be formally defined as follows: let x be the input sentence, represented as a (*x*_1_, …, *x*_*m*_), and let *x*′ be a “blank” baseline input. We have a trained model *F*, and *F* (*x*)_*n*_ is the output of the model at time step n. The contribution of the *mth* word in *x* to the prediction of *F* (*x*)_*n*_ can be calculated by taking the integral of gradients along the straight line path from *x*′ to the input *x*. In other words, we are measuring how much the prediction at time step *n* changes as we move from the baseline input *x*′ to the actual input *x*, and specifically how much the *mth* word in *x* contributes to this change.

The word importance value of each word in the input is calculated by summing the scalar attributions across the dimensions of the input embeddings. A positive attribution value indicates that the word contributed to the prediction made by the model, while a negative attribution value indicates that it opposed the prediction. In cases of the BERT model, which uses sub-word tokenization to divide rare words into smaller pieces, we can obtain word-level attributions that are more understandable to humans by taking the sub-word with the highest absolute attribution value as the attribution for the entire word.

### 3.4 Validating Word Importance

The assumption to use heat-maps of attribution values over the inputs as explanations is particularly popular for natural language processing. To test the validity of these explanations, “stress tests” can be designed using a method called erasure, where the most or least important parts of the input, as indicated by the explanation, are removed and the model’s prediction is observed for changes [2]. Specifically, we erase the most (or least) important word from the input sentence and measure the resulting model accuracy.

### 3.5 Aggregating word attribution

We aggregate the word attributions across multiple texts for each imaging protocol. Integrated gradient assigns attribution scores to each prediction made on a text segment that is a maximum of 512 sub-words long. We calculate the top 5 words for each imaging protocol by taking the average attribution value for each word across all text for a given imaging protocol, and select the top words as those with the highest average attribution value. We further filter out words that appear in less than 3 texts. A trained radiologist assigned a measure of word importance across all text for a given imaging protocol. This measure was based on a numerical score, with a value of 1 indicating a strong influence on the radiologist’s decision, 0.5 indicating a slight influence, and 0 indicating a neutral influence. For each word, the human word importance score was determined as the average of all word scores across a single image protocol class. These methods were employed to generate lists of the most influential words for each imaging protocol, utilizing both the BERT model and the judgments of the trained radiologist.

## 4 Results and Analysis

The results of our fine-tuning experiment on the BERT model are shown in Table 1. The model’s performance was evaluated using three metrics: precision, recall, and F1 score. The F1 score is a measure of the model’s accuracy, taking into account both the precision and recall of the model. We found that the BERT model had an F1 score of 0.89, which represents a significant improvement over the results of previous studies using other machine learning methods. One such study using deep neural network, random forest algorithm, and k-nearest neighbors (kNN) achieved a F1 scores of only 0.83, 0.81 and 0.76 respectively[14].

For our dataset, we also measured the weighted average F1 scores of the traditional machine learning models: XGBoost achieved an F1 score of 0.84, RF scored 0.77, KNN obtained 0.70, and the DNN yielded an F1 score of 0.85. These results are comparable to the performance of existing studies. Overall, the results of our experiment demonstrate the superior performance of the pre-trained BERT model compared to non-Transformer based approaches. The BERT model was able to achieve a higher level of accuracy, as indicated by the higher F1 score, and outperformed other methods in this task.

### 4.1 Word Importance

The attribution scores assigned to individual words by the integrated gradients are intended to reflect the influence of those words on the model’s decisions.

To verify the validity of these attribution scores, we conducted a “stress test” using a technique called erasure. This involved systematically removing the most and least important words from the input text, and measuring the resulting impact on the performance of the BERT model. The results of this stress test are shown in Figure 2. We can see that the removal of the least important words had a relatively small effect on the model’s performance, causing a decline in the F1 score from 0.89 to 0.86. In contrast, the removal of the most important words had a much more significant impact, with the F1 score dropping sharply from 0.89 to 0.62 when the topmost important words was removed. Each subsequent removal of the most important words also resulted in a decremental drop in the F1 score.

**Figure 1.**
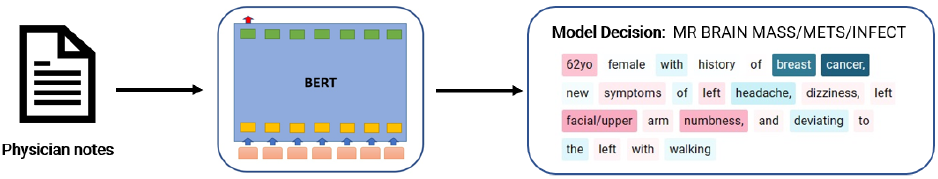
A proposed system in which physician notes are used as input to a model. The output of the model is an imaging protocol, as well as an explanation of the process by which the protocol was determined. This system aims to provide a more efficient and accurate method for determining appropriate imaging protocols, while also offering insight into the decision-making process of the model. By incorporating an explainability component, the proposed system has the potential to enhance trust and understanding in the use of machine learning for medical image protocol assignment.

**Figure 2.**
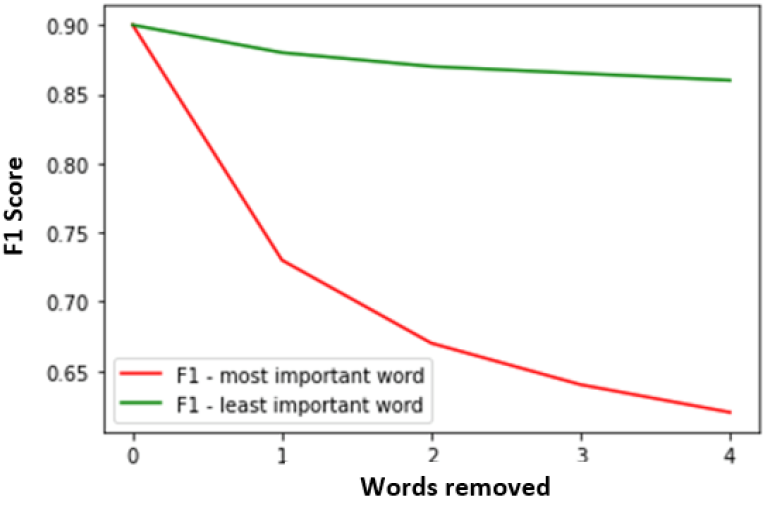
Model performance after step-wise removal of the 4 most important words and the 4 least important words from the text prompt. The results show that the least important words are less likely to degrade model performance while the most important words substantially degrade the performance.

**Figure 3.**
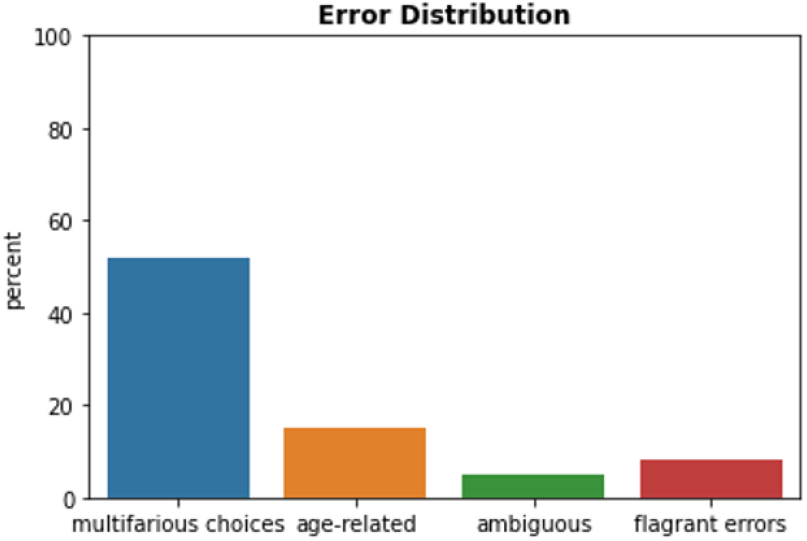
The bar plot decomposes the mistakes into four categories: multifarious choices, age-related, ambiguous text, and flagrant errors.

These results provide strong evidence that the attribution scores generated by the integrated gradients method are valid, as they accurately reflect the influence of each word on the model’s performance. The stress test demonstrates that the most important words have a substantial impact on the model’s ability to make accurate predictions, and that the words with the highest attribution scores are particularly influential in the model’s decision making process.

We aggregate word attribution scores for each image protocol and investigate the difference in the word importance ranks of BERT and those of a radiologist (figure 4). Both human (trained radiologist) and BERT picked the words most frequently mentioned in the indications for brain mass workup. Meningioma is the most common type of brain tumor and lung cancer is the most common cause of brain metastases. Mets is a very commonly used shorthand for metastases. Both human and BERT picked up words suggesting a history of treatment for brain tumors, human picked ‘cyberknife’, while BERT picked ‘post, stereo, treatment’. ‘Rule’ and ‘date’ favored by BERT are most likely due to bias.

**Figure 4.**
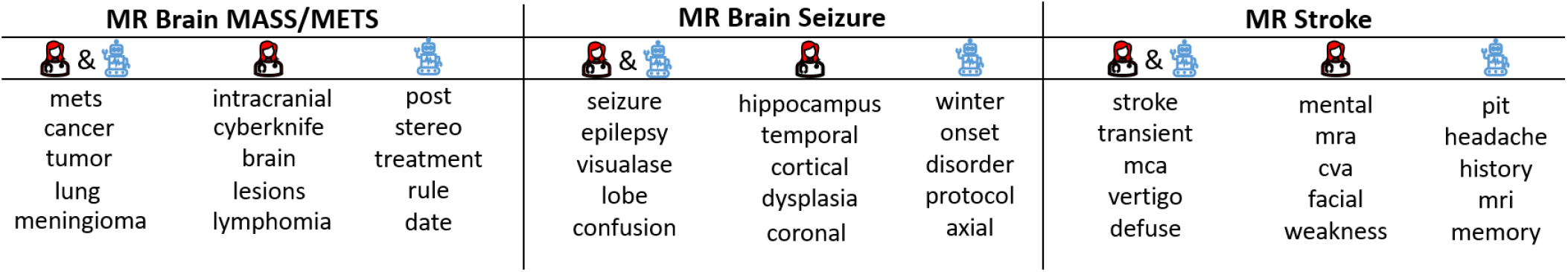
Top 5 words where human (trained radiologist) and BERT agree or disagree for 3 selected protocols. Human & robot are words both human and BERT agree are important. Human only are words with high human importance but low BERT importance. Robot only are words with high BERT importance but low human importance.

Seizure and epilepsy (a condition with prolonged or repetitive seizures) are obviously important for the seizure protocol, both human and BERT agreed. They also consider ‘visualase’, which is an ablation technique for treating seizures, important. BERT did not recognize the specific anatomic structures (hippocampus, temporal lobe) and specialized medial term that are considered important for humans. Instead BERT was biased by some non-specific words.

The top 5 words in agreement for stroke protocol are indeed critical, specific, and frequently used. Again BERT was biased by a few generic words, and failed to recognize words that describe the symptoms of stroke or the medical acronym for stroke (‘cva’).

Furthermore we examine individual texts and their word attribution values to assess the model’s understanding of language in the context of human anatomy and pathology. Figure 5 presents a physician’s text alongside the model’s corresponding word attribution values. In the first example, the model places emphasis on the patient’s history of breast cancer and a headache. In older patients, headaches can often indicate the presence of a brain tumor, and cancer can spread from the breast to the brain, leading to brain metastasis. Despite the presence of symptoms such as dizziness, facial, and numbness, which suggest the possibility of a stroke, the model de-emphasizes these words and correctly determines that brain metastasis is the most likely cause, given the patient’s history of breast cancer and a headache. In the second example, we see a case where the model makes an incorrect decision. The mention of possible edema on a computerized tomography scan suggests the possibility of a brain tumor. Additionally, the model ignores the age of the patient, which is relevant because for patients over the age of 50, seizures are often caused by brain tumors. While an MRI to diagnose brain seizure is plausible, the reasons described indicate that an MRI to diagnose brain metastasis is generally more likely in this case.

**Figure 5.**
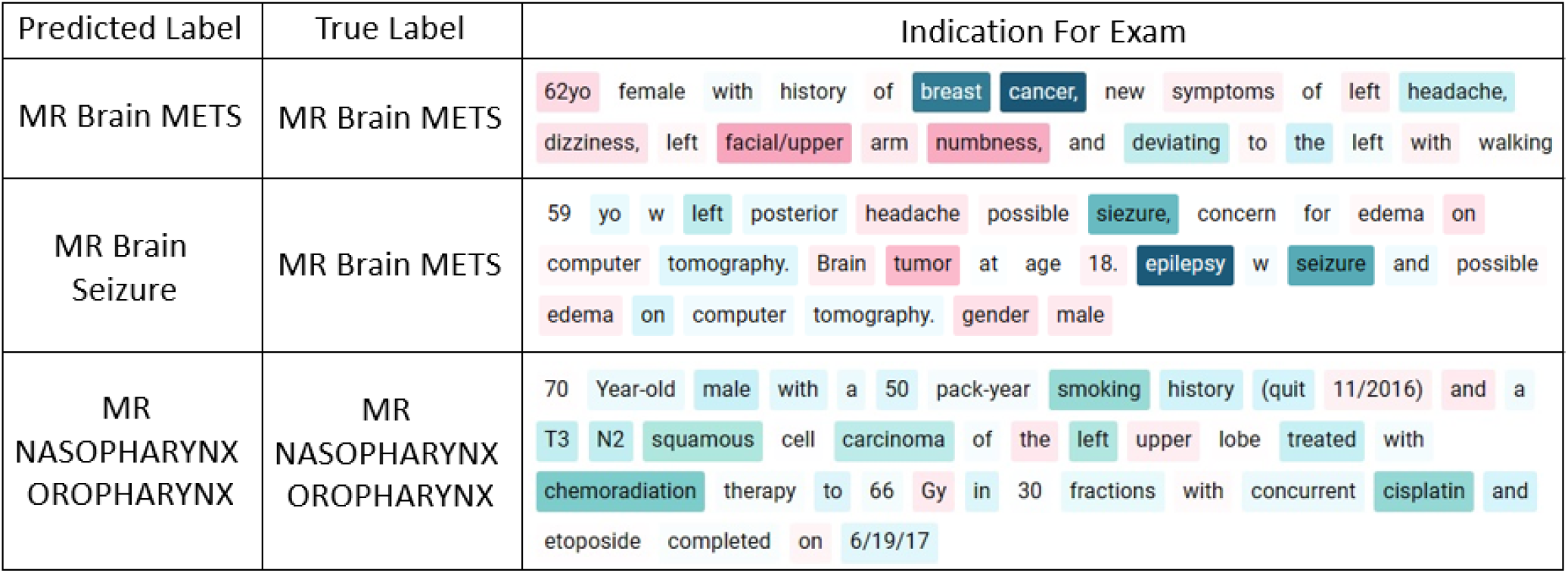
Selected samples from the dataset. The indication for the exam is provided by the ordering physician, which briefly summarizes the symptoms, relevant medical history, and the medical questions. The ‘true label’ is the protocol, assigned manually by a trained radiologist, that is most suitable for the indication. The ‘predicted label’ is the protocol predicted by the AI model.

### 4.2 Error Analysis

In order to understand the errors made by our fine-tuned BERT model on the test set, we conducted an analysis of the model’s explanations and looked for any systematic patterns in the mistakes. Our analysis identified four broad categories of errors: (1) multifarious choices, (2) age-related results, (3) ambiguous entries, and (4) flagrant errors.

The most common type of mistake occurred when the clinical question was too complex or broad, with multiple clinical questions, regions of interest, or complex medical histories. In these cases, there may be multiple valid imaging protocols, and the model struggled to select the most appropriate one. This accounted for 52% of the errors in the test set.

Errors in the second category, age-related results, occurred when the model failed to consider the age of the patient in its prediction. For example, the best protocol for a patient with intracranial hemorrhage may vary depending on their age group. This category accounted for 15% of the errors in the test set. Errors in the third category, ambiguous entries, occurred when the model was unable to make a prediction due to ambiguous or esoteric language in the input text. This could include stems that were too rare or cryptic, or protocols that could not be designated to ambiguous stems. This category accounted for 5% of the errors in the test set. Finally, flagrant errors, the fourth category, occurred when the model made a wrong prediction or the order of word importance did not make sense for the prediction. This category accounted for 28% of the errors in the test set.

Overall, the largest issue for the model was its difficulty in understanding the hierarchical ordering of protocols. This accounted for 52% of the errors in the test set, and will require further work to address before the model can be used in a clinical setting. Another issue was the model’s partial capture of important regions of the input text, which accounted for 15% of the errors. This may be due to biases or limitations in the training data, and will also require further work to address. By understanding the patterns of errors made by the model, we can begin to identify areas for improvement and fine-tune the model to achieve even better performance.

## 5 Discussion

Protocoling is a crucial task for radiologists to ensure that the appropriate sequences are acquired in response to clinical questions. However, manual protocoling can be time-consuming, disruptive, and prone to errors. In recent years, the volume of radiologic orders has increased, making protocoling an increasingly costly burden. To address these challenges, we utilized a large pre-trained language model that was fine-tuned by training it with a large dataset of radiologic orders. This allowed the model to learn medical terminology and accurately process orders, which frequently contain typos, acronyms, and grammatical errors, and are often written in shorthand using specialized medical terminology.

Furthermore, in response to the increasing demand for ‘explainable AI’, we investigated the decision-making process of our model. We evaluated the model’s ability to provide explanations of its decision based on ‘word importance’. Model explanation techniques were applied to estimate the importance of each word within the text of each radiologic order. This allowed us to delve into the model’s decision-making process and determine whether it was making correct predictions for the right reasons, as well as to identify the root causes of any mistakes. Our results indicate that the BERT model is able to identify relevant words that are highly indicative of the target protocol.

Our error analysis revealed that the model struggled most with understanding complex indications involving multiple clinical questions, leading to incorrect protocol selection in some cases. For example, the model may have difficulty distinguishing between protocols for a patient with acute neurologic deficits after brain tumor resection, as it may not fully comprehend the hierarchical ordering of protocols. Furthermore, we identified that approximately 15% of the model’s mistakes were due to insufficient capture of important regions of the input text. This could be due to various factors such as bias in the training data or limited examples of certain edge cases.

Overall, the utilization of integrated gradients in our analysis has provided valuable insights into the model’s decision-making process compared to that of a trained radiologist. This information was used to evaluate the strengths and weaknesses of the model, and will be instrumental in making the model more robust and trustworthy before its application in clinical settings.

## 6 Limitation

There are several limitations to consider in the context of this study. First, our dataset comprised of neuroradiologic orders from a single center, and thus may be limited in its representation of the racial, social, and ethnic diversity of other regions. Validation with datasets from different institutions is necessary to more accurately compare the model’s performance. Additionally, we limited the number of protocols to the ten most commonly used protocols in this study, which may not fully capture the breadth of protocols used in clinical practice. The data was collected from routine clinical work, which means that protocols were assigned by multiple radiologists with varying levels of experience, potentially leading to inter-operator variability. While the dataset is relatively large at over 80,000 entries, it is possible that additional data could further improve model performance.

Additionally, it is important to note that there may be significant variations in the importance of certain words when considering the perspectives of different radiologists. In this study, we were constrained to a single radiologist when evaluating word-level agreement with BERT. However, in future studies, it would be beneficial to evaluate word importance from the perspectives of a diverse group of radiologists to achieve more robust results.

## 7 Related Work

Previous work has been done using classification models to predict imaging protocol from a physician’s notes using machine learning techniques such as SVM, Random Forests, and Gradient Boosted Machine [5, 6]. More recently, a deep neural network approach was used to automate radiological protocols which showed a slight boost over kNN and random forests [14]. However, these models are limited by the size of the model and the use of classical word embeddings which don’t provide deep contextual word embeddings [27]. To date, there has been no research on explainable medical text for image protocol classification tasks or on the decision-making process of these models to identify potential systematic errors that may need to be addressed.

Recently bidirectional RNN’s and transformers have improved text representation to be sensitive to its local context in a sentence and optimized for specific tasks by using a self-attention mechanism to help embed the context of each word [25]. Large language models such as BERT [9] and ELMo [20] have been shown to provide substantial performance improvements for language modeling and text classification. We hypothesize that the use of context-dependent token embeddings will provide a substantial improvement for medical text classification and model interpretation. While there has been recent work evaluating large pretrained models for specialized tasks such as legal contract review [13], to the best of our knowledge, this paper is the first to evaluate how these models will perform on this specialized medical text which poses different challenges.

Furthermore, in the case of high stake applications, both accuracy and trust are necessary for the adoption of the model’s decisions. Recent studies have focused on incorporating model explanations to improve trust [17]. Explainable models have been developed to visualize word importance and attention layers [11]. This has provided researchers with insight into understanding the model’s decisions [12]. However, to the best of our knowledge, no other group has attempted to evaluate if machine learning models can provide valid explanations for specialized medical texts.

## 8 Conclusion

In this study, we demonstrate state-of-the-art performance for the radiologic protocol classification task and provide a better understanding of how natural language processing (NLP) models make decisions in the medical domain. Using a large dataset of over 80,000 entries annotated by medical experts, we evaluated a pretrained BERT model and found that it significantly outperformed existing machine learning methods. We showed that BERT is able to identify relevant words that are highly indicative of the target protocol. The differences in BERT and human word importance were driven by BERT not recognizing specific anatomic structures and specialized medial terms that are important for humans. Furthermore, our analysis of the errors revealed that the largest source of errors was due to the model’s difficulty in understanding the hierarchy of protocol assignments, while the third largest contributor was potential limitations or biases in the dataset.

Overall, our findings demonstrate that BERT can provide valuable insight into its decision making process for specialized medical tasks. This insight is valuable in understanding the error profile of the model. Understanding BERT’s decision making process is a necessary stop to deploying it in a real life clinical environment.

## Data Availability

All data produced in the present work are contained in the manuscript

## 10 AUTHOR CONTRIBUTIONS

ET and ST conceived of the research study. ST contributed toward the design, implementation and evaluation of machine learning models. ET curated the dataset and evaluated the model’s errors. ET, MM ST managed the project vision and implementation along with writing of the manuscript.

## 11 DATA AVAILABILITY

The datasets utilized during this study are not publicly available due to reasonable privacy and security concerns. The data is not easily redistributable to researchers other than those engaged in the Institutional Review Board-approved research collaborations with Stanford University.

